# Grey Matter Volume and Fractional Anisotropy as Correlates of Cognitive Improvement in Traumatic Brain Injury Over a 6-Month Period

**DOI:** 10.1101/2024.10.24.24315709

**Authors:** Ben Zhang, Niko Fulmer, Sarah Dunn, Zhong Sheng Zheng, Caroline Schnakers, Emily R. Rosario

## Abstract

**Objective:** In this study we explored how neuroimaging and blood biomarkers relate to cognitive recovery in traumatic brain injury (TBI) patients.

**Methods:** Sixteen participants with moderate to severe traumatic brain injury (TBI) were enrolled, with blood samples, MRI, and diffusion tensor imaging (DTI) collected at enrollment and six months. The Repeatable Battery for the Assessment of Neuropsychological Status (RBANS), Disability Rating Scale, and Montreal Cognitive Assessment (MoCA) were also administered at both time points to evaluate neuropsychological and functional outcomes.

**Results:** Fractional anisotropy (FA) in the genu (r_s_ = 0.937, p = 0.002) and splenium of the corpus callosum (r_s_ = 0.955, p < 0.001) was strongly correlated with changes in RBANS - Attention scores. Fornix FA was correlated with changes in RBANS - Total (r_s_ = 0.928, p = 0.008), and left tapetum FA was correlated with changes in RBANS - Visuospatial scores (r_s_ = 0.964, p < 0.001). Right temporal fusiform cortex grey matter (GM) volume was correlated with changes in RBANS - Attention scores (r_s_ = 0.975, p = 0.005). Blood biomarkers did not show significance.

**Conclusion:** Imaging markers like FA and GM volume appear to help predict cognitive recovery in TBI, supporting the potential use of neuroimaging to guide rehabilitation strategies.

## Introduction

Traumatic brain injury (TBI) is a major cause of long-term disability globally, leaving survivors with a range of cognitive, emotional, and physical challenges. Due to the significant variability in individual recovery, identifying reliable biomarkers to predict recovery trajectories has become a critical focus of research. Understanding which structural and biological factors, such as brain volume changes and molecular markers, influence cognitive recovery is key to improving prognosis, guiding personalized rehabilitation strategies, and ultimately enhancing quality of life for TBI survivors (2).

Patients often undergo routine MRI scans as part of their initial evaluation after being admitted to the hospital following the acute phase of TBI (3). These scans provide high-resolution T1-weighted images, allowing for the assessment of grey matter volume. Additionally, diffusion tensor imaging (DTI), though less widely available, can measure white matter integrity, which is particularly vulnerable to damage during traumatic events. White matter tracts are crucial for communication between different brain regions, and are particularly susceptible to injury during traumatic events (4). Using DTI metrics like fractional anisotropy (FA) we are able to obtain detailed information regarding their microstructural integrity. Computing FA, alongside measurements of grey matter volumes in regions of the cerebral cortex, allows for the identification of potentially significant correlates of cognitive recovery (5).

Blood-based markers serve as another variable for monitoring physiological processes of interest. Measures such as Brain-Derived Neurotrophic Factor (BDNF), Interferon Gamma (IFN-γ) and C-Reactive Protein (CRP) are among many which have been implicated in the pathophysiology of TBI (6,7,54). For example, CRP is a measure of systemic inflammation that has been associated with less desired cognitive outcomes (8).

This study aims to explore the prognostic value of baseline neuroimaging and blood markers in predicting cognitive recovery in TBI patients. By analyzing correlations between baseline white matter integrity, grey matter volume, and cognitive changes alongside blood data, we aim to identify biomarkers that can reliably estimate recovery trajectories. Early recognition of patients at risk for poorer outcomes could significantly improve the management and rehabilitation of their treatment, and ultimately result in better long-term recovery.

## Participants

This study included 16 participants (mean age at time of injury: 41.02 years, SD = 13.83) with moderate to severe TBI who were previously enrolled from acute and subacute hospital settings (mean time since injury: 22.27 months, SD = 39.74). Inclusion criteria required participants to have a diagnosis of moderate to severe TBI, be fluent English speakers, have intact motor use of their dominant hand, and have no receptive or expressive language impairments, including aphasia. Participants were excluded if they were under the age of 18 or over the age of 65 years. Additionally, individuals who had claustrophobia or metallic implants that would make MRI assessments unsafe were also excluded from the study. Demographic and clinical data of the enrolled participants are detailed in Table 1. The study was conducted following approval from the Institutional Review Boards at Casa Colina Hospital and Pomona Valley Hospital. All participants provided written informed consent prior to their inclusion in the study.

**Table 1.** Demographics.

| Variable | Mean (SD) |
| --- | --- |
| Age at Time of Injury | 41.02 years (13.83) |
| Months Post Injury | 22.27 (39.74) |
| Male / Female / Unsure | 12 / 3 / 1 |
| Hispanic / Non-Hispanic / Unsure | 3 / 12 / 1 |

## Blood Data Acquisition

Blood samples were drawn from participants via venipuncture and collected in serum separator tubes at two protocol timepoints (study enrollment and at the 6 month follow up). Blood draws occurred at various times throughout the day and tubes were allowed to clot at room temperature for 30 minutes before centrifugation at 1,500 x g for 10 minutes. The serum was then aliquoted and stored at -80°C until further analysis. On the initial freeze-thaw cycle, biomarker levels were quantified for IFN-γ, IL-6, IL-8, TNF-α, CRP, IL-10, and BDNF using the EllaTM platform, a SimplePlexTM high sensitivity automated microfluidic multiplex immunoassay system using cartridges to quantify in triplicate, various analyte signals through laser camera capture (Protein Simple, Biotechne).

## MRI Acquisition

MRI data was acquired using a 3T Siemens Magnetom Verio scanner. A T1-weighted Magnetization Prepared Rapid Gradient Echo (MPRAGE) sequence was used for the high resolution 3-D anatomical scan. It had the following parameters: repetition time (TR) of 2300 ms, echo time (TE) of 2 ms, a flip angle of 9°, a field of view (FOV) of 230 mm, and a slice thickness of 1 mm without any gaps. The scan included 160 slices with a matrix size of 224 × 224. Diffusion-weighted imaging (DWI) was conducted with the following settings: TR of 10900 ms, TE of 95 ms, slice thickness of 2 mm with no gaps, 82 slices, and a matrix size of 122 × 122. Diffusion-sensitizing gradients were applied in 64 different non-collinear directions with a b-value of 1000 s/mm^2^. Additionally, five images without diffusion weighting (b = 0) were captured.

## T1/DTI Preprocessing & Analysis

First, T1-weighted images underwent bias-field correction using FSL tools (available at http://www.fmrib.ox.ac.uk/fsl). Brain extraction was then performed using the Optimized Brain Extraction for Pathological Brains (optiBET) technique, and grey matter segmentation was carried out with FMRIB’s Automated Segmentation Tool (FAST) (9). We took a region of interest (ROI) approach with labels from the Harvard-Oxford cortical structural atlas (available at https://neurovault.org/collections/262/), focusing on temporo-cortical regions given the substantial evidence highlighting the relationship between TBI severity and the temporal lobe (10-16).

Seventeen ROIs were selected (Figure 1) : temporal pole, anterior and posterior divisions of the superior temporal gyrus, anterior and posterior divisions of the middle temporal gyrus, temporooccipital middle temporal gyrus, anterior and posterior divisions of the inferior temporal gyrus, temporooccipital inferior temporal gyrus, anterior and posterior divisions of the parahippocampal gyrus, anterior and posterior divisions of the temporal fusiform cortex, temporal occipital fusiform cortex, planum polare, heschl’s gyrus, and planum temporale.

**Figure 1.**
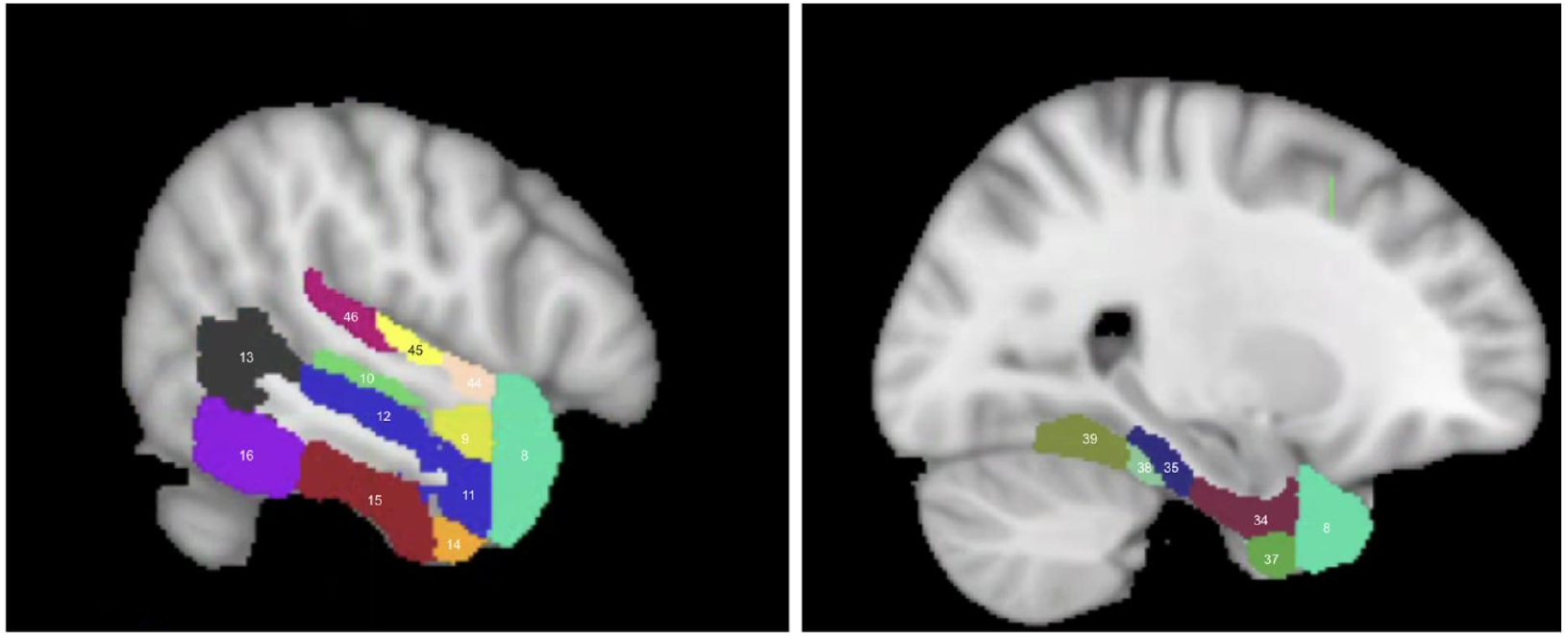
Temporal Cortical Grey Matter Regions of Interest. Regions are labeled as thresholded on Harvard Oxford Cortical Structural Atlas: (8) temporal pole, (9) superior temporal gyrus - anterior division, (10) superior temporal gyrus - posterior division, (11) middle temporal gyrus - anterior division, (12) middle temporal gyrus - posterior division, (13) middle temporal gyrus - temporooccipital part, (14) inferior temporal gyrus - anterior division, (15) inferior temporal gyrus - posterior division, (16) inferior temporal gyrus - temporooccipital art, (34) parahippocampal gyrus - anterior division, (35) parahippocampal gyrus - posterior division, (37) temporal fusiform cortex - anterior division, (38) temporal fusiform cortex - posterior division, (39) temporal occipital fusiform cortex, (44) planum polare, (45) Heschl’s gyrus, (46) planum temporale

In order to align the cortical ROIs with the patients’ structural data, we performed nonlinear registration between the standard MNI space and each patient’s structural space. Grey matter volume was then calculated for each ROI after they were manually inspected for accuracy.

For preprocessing the DWI data, we utilized DTIPrep to identify and remove volumes with artifacts. This step included corrections for eddy currents and head motion through affine registration to the mean b=0 reference image. The Brain Extraction Tool (BET) was then used to strip the skull from the b=0 image. At each voxel in the brain, diffusion tensors were computed using the diffusion toolbox in FSL, and FA maps were generated. We applied an ROI approach to this as well, using labels from the John Hopkins University (JHU) white matter atlas (available at http://cmrm.med.jhmi.edu).

Alongside the relationship with the temporal lobe, literature relating to DTI suggests a strong connection between interhemispheric white matter and cognition following TBI. Hence, we opted to examine not only temporal white matter tracts, but those of the corpus callosum as well (17-20). The following ROIs were selected (Figures 2 & 3) : uncinate fasciculus, cingulum, sagittal stratum, superior longitudinal fasciculus, stria terminalis, fornix, posterior thalamic radiation, genu of corpus callosum, body of corpus callosum, and splenium of corpus callosum, and tapetum.

**Figure 2.**
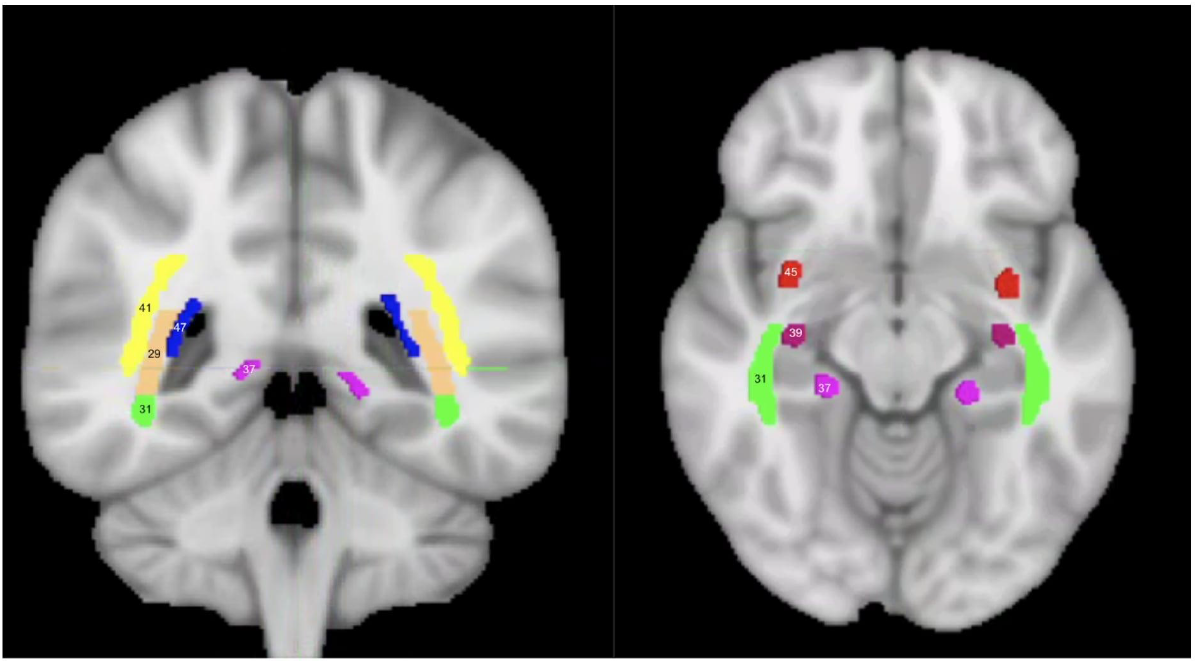
Temporal White Matter Regions of Interest. Regions are labeled as thresholded on John Hopkins University White Matter Atlas : (29) posterior thalamic radiation, (31) sagittal Stratum, (37) cingulum (hippocampus), (39) stria terminalis, (41) superior longitudinal fasciculus, (45) uncinate fasciculus, (47) tapetum

**Figure 3.**
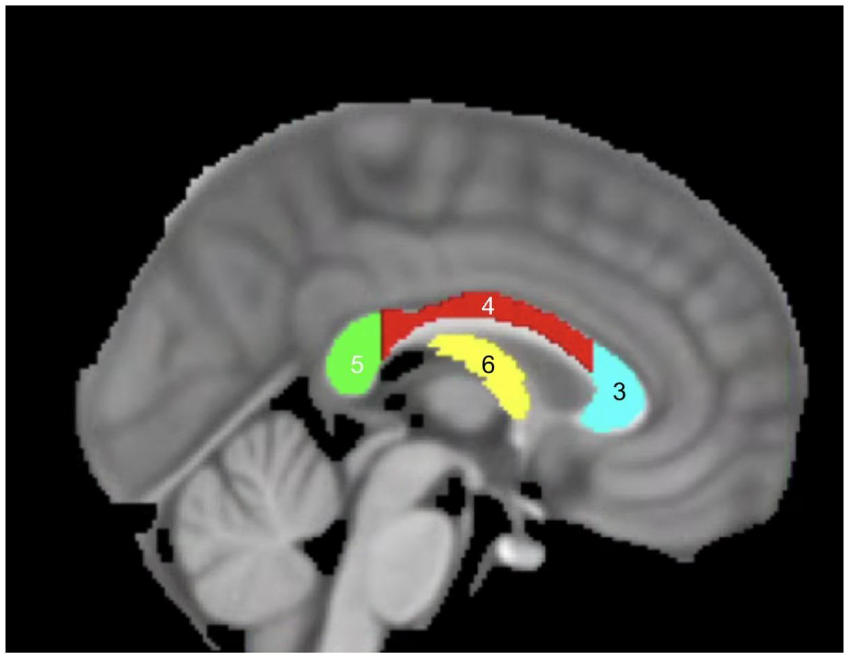
Corpus Callosum & Fornix White Matter Regions of Interest. Regions are labeled as thresholded on John Hopkins University White Matter Atlas : (3) genu of corpus callosum, (4) body of corpus callosum, (5) splenium of corpus callosum, (6) fornix

A similar method to the T1 data was used to align the standard white matter tract ROIs with the patients’ diffusion data. Nonlinear registration between the standard MNI space (FMRIB58_FA) and each patient’s diffusion space was done, then each ROI was visually inspected for accuracy and manually corrected if necessary before data analysis. To ensure that only white matter voxels were included, a whole-brain white matter segmentation mask was generated for each patient using the processed T1-weighted image and then intersected with the ROIs. Finally, the average FA value for each ROI was calculated for each patient.

## Procedure / Cognitive Assessments

We evaluated the recovery trajectory of TBI patients over a six-month period following their injuries. Data was collected immediately following study enrollment, and then after 6 months during a dedicated one-week assessment period. During each of these assessment weeks participants underwent a series of structured evaluations which were designed to provide a comprehensive overview of their recovery progress. Demographic data was collected for each participant at the beginning of their enrollment in the study.

Evaluations conducted at each interval included blood sample collections, neuropsychological assessments, and magnetic resonance imaging. The following neuropsychological evaluations were administered by trained clinicians. Montreal Cognitive Assessment (MoCA), consisting of 30 total points : visuospatial abilities (5 points), confrontation naming (3 points), short term memory (5 points), attention (6 points), language (3 points), abstraction (2 points), and orientation (6 points). The Repeatable Battery for the Assessment of Neuropsychological Status (RBANS), with an index score range of 40-160, contained list learning (40 points), story memory (24 points), figure copying (20 points), line orientation (20 points), picture naming (10 points), semantic fluency (40 points), digit span (16 points), list recall (10 points), list recognition (20 points), story recall (12 points), and figure recall (20 points). Total scores were converted to index scores. The Disability Rating Scale (DRS) has a maximum score of 29, correlating with an extreme vegetative state - a person without disability would have a score of 0. It measures eye opening (3 points), communication ability (4 points), motor response (5 points), feeding awareness (3 points), toileting awareness (3 points), grooming awareness (3 points), level of functioning (5 points), employability (3 points).

## Statistical Analysis

Statistical analyses were conducted using Jeffrey’s Amazing Statistics Program (JASP) and RStudio with a *p*-value of 0.05 for significance (21,22). Due to the non-parametric nature of the data, Spearman’s rank-order correlation analyses were used to assess the relationships between initial blood and neuroimaging variables with changes in cognitive scores. Prior to conducting these analyses, we tested whether age and time since injury were significantly correlated with the variables of interest. Both variables were found not to have significant correlations, and as such they were not included as covariates in the final analyses. The Benjamini-Hochberg method (FDR = 0.05) was utilized for multiple comparisons correction.

Non-parametric (Wilcoxon) paired sample T-tests were conducted on blood data, RBANS, DRS, and MoCA scores in order to identify significant changes throughout the study.

## Results

After applying multiple comparisons corrections, a significant correlation was found between the initial grey matter volume of the posterior division of the right temporal fusiform cortex and 6 month change in RBANS - Attention scores (r_s_ = 0.975, p = 0.005).

FA in the fornix was correlated with change in RBANS - Total (r_s_ = 0.928, p = 0.008). Initial white matter integrity in corpus callosum white matter tracts also had significant associations with cognitive outcomes. Specifically, change in the RBANS - Attention score was correlated with baseline FA in the genu of the corpus callosum (r_s_ = 0.937, p = 0.002) as well as the splenium of the corpus callosum (r_s_ = 0.955, p < 0.001). FA in the left tapetum was correlated with change in RBANS - Visuospatial (r_s_ = 0.964, p < 0.001).

Without passing multiple comparisons corrections, FA in the body of corpus callosum, splenium of corpus callosum, right tapetum as well as in the right posterior thalamic radiation exhibited trending correlations with change in RBANS - Visuospatial/Construction. See Tables 5-6 for results, and Figure 4 for scatterplots of significant correlations.

None of the examined blood-based biomarkers: IFN-γ, IL-6, IL-8, TNF-α, CRP, IL-10, and BDNF, were significantly correlated with changes in cognitive scores. T-tests examining change from study enrollment to 6 months for the blood biomarkers did not yield significant results. However, due to blood markers in this small cohort, these findings may not fully capture the potential relationships and changes, underscoring the need for larger datasets to confirm these results.

There was no significant change in MoCA, DRS, or RBANS from study enrollment to the 6 month follow up. Refer to Tables 2-3 for specific information regarding blood and cognitive scores at both timepoints.

**Table 2.** Baseline and 6-Month Cognitive Scores.

| Variable | Baseline Mean (SD) | 6 Month Mean (SD) |
| --- | --- | --- |
| Montreal Cognitive Assessment (MoCA) | 21.28 (4.61), n = 14 | 22.33 (3.87), n = 9 |
| Disability Rating Scale (DRS) | 3.67 (2.58), n = 15 | 2.9 (2.08), n = 9 |
| Repeatable Battery for the Assessment of Neuropsychological Status (RBANS) | 67.07 (10.54), n = 13 | 74.75 (13.98), n = 8 |

**Table 3.** Baseline and 6-Month Blood Markers.

| Variable | Baseline Mean (SD) | 6 Month Mean (SD) |
| --- | --- | --- |
| IFNγ | 0.873pg/ml (0.42), n = 10 | 0.675 pg/ml (0.27), n = 8 |
| IL-6 | 2.379 pg/ml (1.41), n = 11 | 1.859 pg/ml (0.91), n = 8 |
| IL-8 | 11.931 pg/ml (7.54), n = 12 | 14.996 pg/ml (6.12), n = 10 |
| TNF-α | 10.67 pg/ml (3.63), n = 12 | 12.712 pg/ml (3.08), n = 10 |
| CRP | 2596857 pg/ml (3059291), n = 8 | 1488985 pg/ml (2282827), n = 8 |
| IL-10 | 2.1 pg/ml (1.35), n = 7 | 2.024 pg/ml (0.94), n = 5 |
| BDNF | 4335.27 pg/ml (3992.6), n = 11 | 9988.8 pg/ml (3745.9), n = 10 |

## Discussion

Our study aimed to explore the various biomarkers of recovery from traumatic brain injury by investigating the relationship between baseline variables and change in cognitive scores over a 6-month period. Our findings can be summarized as follows : 1) Baseline FA in temporal and interhemispheric white matter tracts were positively correlated with changes in various cognitive scores; 2) Baseline grey matter volume in the right temporal fusiform cortex was positively correlated with improvements in attention; 3) Blood biomarkers did not have any relation with cognition, however more data is needed to draw conclusions.

The temporal fusiform cortex is the outer layer of grey matter on the fusiform gyrus that is located on the temporal lobe (23). It is strategically positioned within the ventral visual stream, which allows it to serve as a key structure in high-level visual processing. It also functions in object recognition and the processing of complex visual stimuli, such as faces (24-26). Higher grey matter volume in this region could positively contribute to change in RBANS - Attention in several ways. First, the temporal fusiform cortex seems to have a direct relationship with attentive ability via the aforementioned visual processing. Increased grey matter in the region could also indicate better structural preservation of neural areas critical for visual processing and attention, which could provide a stronger foundation for recovery over time (27). Studies have also shown that neuroplasticity can be enhanced via increased use of relevant brain regions - indicating a possibility of heightened attentive ability leading to increased improvements in attention (28,29).

FA has been found to be a proxy for the integrity of white matter microstructures such as axons and myelin. Following brain injuries, lower levels of FA are usually associated with worse cognitive and behavioral outcomes (18,30). Relevantly, the fornix is a C-shaped bundle of white matter fibers which act as the primary output tract of the hippocampus. It includes fibers from various hippocampal subregions such as the dentate gyrus and subiculum, playing a crucial role in memory consolidation and retrieval (23, 31, 32). Our findings of a positive association between white matter integrity of the fornix and changes in RBANS - Total scores over a six month period could be explained by higher FA indicating a more efficient pathway for information transfer between short and long-term memory (33,34). Consequently, we infer a relation to faster learning and better information retention over the recovery period. This would influence multiple cognitive domains assessed by RBANS, and could also facilitate neuroplastic changes over time (35).

The significant correlations between initial FA in the genu and splenium of corpus callosum with RBANS - Attention further underscore the importance of white matter microstructure in cognitive recovery following TBI. The genu, consisting of white matter fibers that connect the prefrontal cortices, is involved in executive functions like attentional control and working memory (36-38). The splenium connects the temporal and occipital regions of both hemispheres, playing a large role in the transfer of visuospatial processing and sensory information between lobes (36, 39, 40).Together the increased FA in these two areas provide a robust foundation for attention-based cognitive recovery and most likely allow for more efficient interhemispheric information transfer. It could also support better top-down control of attention usage in regards to visuospatial tasks, which could result in faster and more accurate shifting between stimuli. This would reflect more cognitive flexibility in neuropsychological testing. The improved efficient interhemispheric communication from these regions could improve the redistribution of cognitive load across both hemispheres, potentially enhancing capacity for adaptive reorganization following TBI as well (41-45).

The left tapetum, a white matter tract that runs along the lateral aspect of the corpus callosum adjacent to the optic radiation, was associated with improvements in RBANS - Visuospatial scores. This association is likely due to its anatomy as it contains fibers from both the splenium and body of corpus callosum (46). It may play a role in visual processing similar to that of the splenium, as well as contribute to interhemispheric processing for visual and spatial information. Alternatively, this correlation could be caused by the challenges of delineating the tapetum from the optic radiations. As registration of the JHU white matter atlas between standard MNI space and subjects’ native diffusion space was necessary for delineation, noise may have been introduced into the measurements. This process could have potentially led to slight misalignments in the delineation of closely adjacent structures (47, 48).Consequently, it is possible that the ROI used to measure the tapetum’s FA may have inadvertently included some of the white matter voxels from the optic radiation. The optic radiation is crucial for the transmission of visual information from the thalamus to the visual cortex, so it could provide a plausible explanation for the observed correlation with visuospatial scores (49, 50). Future exploration should look to disentangle the contributions of the optic radiation and tapetum in visuospatial recovery.

No significant effect was found between blood markers at enrollment and changes in cognitive scores. However, we are reluctant to attribute this to lack of relationship, as current literature points towards an association between many blood markers with recovery following stroke and brain injury (6, 7, 54). Multiple participant’s blood concentrations were out of range of measurement, leading to a small sample size of blood data.

Overall, these findings illuminate the interplay between the temporal areas of the brain and cognitive recovery following TBI. They suggest that participants’ white matter integrity and cortical brain volume play significant roles in the recovery process, reiterating the need for personalized approaches in the rehabilitation of TBI patients.

## Limitations

This study is subject to certain limitations. Its focus on specific regions of interest, while informative, could overlook other critical regions involved in TBI recovery. There is also a lack of exploration of the subcortical brain regions. Future research should aim to explore a broader range of brain regions and incorporate more specific delineation of the optic radiation and tapetum in order to provide a more complete picture of the changes following TBI.

While blood biomarker data did not yield insights into cognitive recovery within our sample, the small number of participants, the blood draw collection time variability, and the complexity of cytokine and chemokine signaling on both systemic and neuro-specific inflammation limits our ability to fully understand its impact. Future studies should be conducted with larger samples of blood markers, with collection at the same time of day, to account for diurnal variation. Studies could also assess the blood biomarker changes over time following trauma through the acute and into the more long term recovery phases post TBI.

Furthermore, the study’s focus on a specific timeframe of baseline to six months post injury may not capture the full trajectory of cognitive recovery, which can continue for years following TBI (51, 52). Each participant joined the study at different lengths of time following their TBI onset, so this could have confounded the analysis with recovery trajectories being different at critical periods following injury (51, 53). A long-term follow up coupled with a consistent time from injury to study enrollment could provide more stability of observed associations.

## Conclusion

This study offers valuable insights into the role of white matter integrity and grey matter volume in cognitive recovery following moderate to severe TBI. Our findings suggest that neuroimaging markers within interhemispheric tracts and temporal regions have significant clinical relevance to the trajectory of cognitive recovery. The observed associations highlight the potential of these markers to serve as indicators of neuroplasticity and recovery potential, and also encourage their integration into personalized rehabilitation strategies. Future research should aim to incorporate additional neuroimaging modalities as well as more brain regions in order to enhance the predictive power of these biomarkers, ultimately improving long-term outcomes for TBI patients.

## Data Availability

All data produced in the present study are available upon reasonable request to the authors

## Funding

### Data availability statement

The data that support the findings of this study are available from the corresponding author, Emily Rosario, upon reasonable request.

## Conflict of interest

The authors declare that the research was conducted in the absence of any commercial or financial relationships that could be construed as a potential conflict of interest.

## Notes

### Competing Interest Statement

The authors have declared no competing interest.

### Funding Statement

This study was funded by the Norris Foundation

### Author Declarations

IRB of Casa Colina Hospital and Centers for Healthcare gave ethical approval for this work.

